# Nature of transmission of Covid19 in India

**DOI:** 10.1101/2020.04.14.20065821

**Authors:** Anushree Roy, Sayan Kar

## Abstract

We examine available data on the number of individuals infected by the Covid-19 virus, across several different states in India, over the period January 30, 2020 to April 10, 2020. It is found that the growth of the number of infected individuals *N*(*t*) can be modeled across different states with a simple linear function *N*(*t*) = *γ* + *αt* beyond the date when reasonable number of individuals were tested (and when a countrywide lockdown was imposed). The slope *α* is different for different states. Following recent work by Notari (arxiv:2003.12417), we then consider the dependency of the *α* for different states on the average maximum and minimum temperatures, the average relative humidity and the population density in each state. It turns out that like other countries, the parameter *α*, which determines the rate of rise of the number of infected individuals, seems to have a weak correlation with the average maximum temperature of the state. In contrast, any significant variation of *α* with humidity or minimum temperature seems absent with almost no meaningful correlation. Expectedly, *α* increases (slightly) with increase in the population density of the states; however, the degree of correlation here too is negligible. These results seem to barely suggest that a natural cause like a hot summer (larger maximum temperatures) may contribute towards reducing the transmission of the virus, though the role of minimum temperature, humidity and population density remains somewhat obscure from the inferences which may be drawn from presently available data.

## I. INTRODUCTION

The ongoing Coronavirus pandemic which has gripped the entire world has been a cause of major existential concern among humans across countries and continents. COVID-19, as the pandemic is technically called, is a highly infectious viral disease which first appeared in Wuhan in China in December 2019. Since then, containing the virus through studies of its transmission mechanism has been a subject of great interest. A central question along these lines is whether local climate (as embodied in the temperature and relative humidity at a given location) on Earth has any role to play in the existence, functional behaviour and transmission of this virus. It is true that apart from temperature or humidity, there could be many other factors as well which could contribute to an answer to this serious question. In 2011, a biology oriented analysis on the role of climatological factors was carried out in [2] for the SARS coronavirus (not the present one). In the last few months or so there has been renewed interest on this question largely in the context of SARS-Cov-2 and its devastating presence across the globe. Analysis done by various groups and individuals appear in [3], [4], [5], [6], [7] and [8]. Much of the analysis in these articles (except [3]) is based on laboratory based biological/medical experimentation and analysis related to the virus.

Let us now try to state how we approach this problem. Our motivation and methods are entirely based on the recent work by Notari [1]. Firstly, we are not *directly* concerned with the medical or biological characteristics of the virus. Our only inputs seem to be the nature in which the number N of infected individuals in a given location changes with time. We have data on this freely available in the Internet. It has been found that the growth of this number *N*(*t*) in places like Italy or China has been exponential, i.e

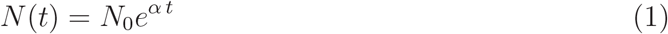

where *N*_0_ and *α* are two parameters. One expects that these parameters will however depend on various factors since they turn out to be different in different places. Assuming a sort of universality across countries and continents we may take the functional form to be the same, though that is also not a necessity.

Since *N*_0_ and *α* depend on various factors, the question arises what could they be. We take *α* as varying with local temperature (following [1]), humidity and also population density. We wish to find out the nature of this variation from the data. Note that, instead of an exponential, any other growing functional form

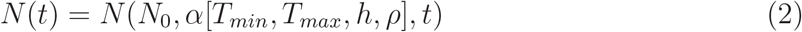

can arise from the nature of the data. For example, Brandenburg proposes a quadratic dependence in a recent analysis [9]. In the above-stated expression (Eqn. 2) *T*_*min*_, *T*_*max*_, *h* and *ρ* denote the minimum temperature, maximum temperature, humidity and population density respectively. As we note later, a linear or quadratic dependency of *α* on the various parameters will be assumed in this work.

Available data on average temperature of a certain location over a certain period of time is easily obtainable from meteorological data. Using such data it is possible to plot *α* as a function of the various quantities (minimum temperature, maximum temperature, humidity and population density) by noting the values of *α* in various states/countries. This type of analysis has been done by various authors [1],[3] very recently using data from different countries. We do a similar analysis here, in the Indian context, for the different states of the Indian Union.

Several groups of Indian scientists in India and abroad have contributed recently on various aspects of Covid19. Some of the work done includes (a) work based on epidemiological models with suggestions on interventions such as lock-down [10] [11], [12],[13], [14]; (b) estimates of the number of infections in Indian hot-spots using fatality data [15] and (c) analysis on the space-time dependence of the outbreak [16].

## II. *N*(*t*) ACROSS DIFFERENT STATES IN INDIA: THE NATURE OF THE VARIATION

Our first step is to plot the data on the number of infected individuals as a function of time. Figure 1 plots the number *N* of diagnosed COVID-19 cases in 15 different states of India since March 02, 2020. We chose the states, wherein the numbers of cases are above 60 till April 10, 2020. The data source is the web portal of the Ministry of Health and Family Welfare, Government of India [17]. The first case was reported from Kerala on January 30, 2020. The number rose to 3 in February. For clarity of the plot, we do not show these two initial data points from Kerala in Figure 1. The next place which got affected was Delhi. The first report from Delhi came on March 02, 2020. From the available data we find the following:

**FIG. 1.**
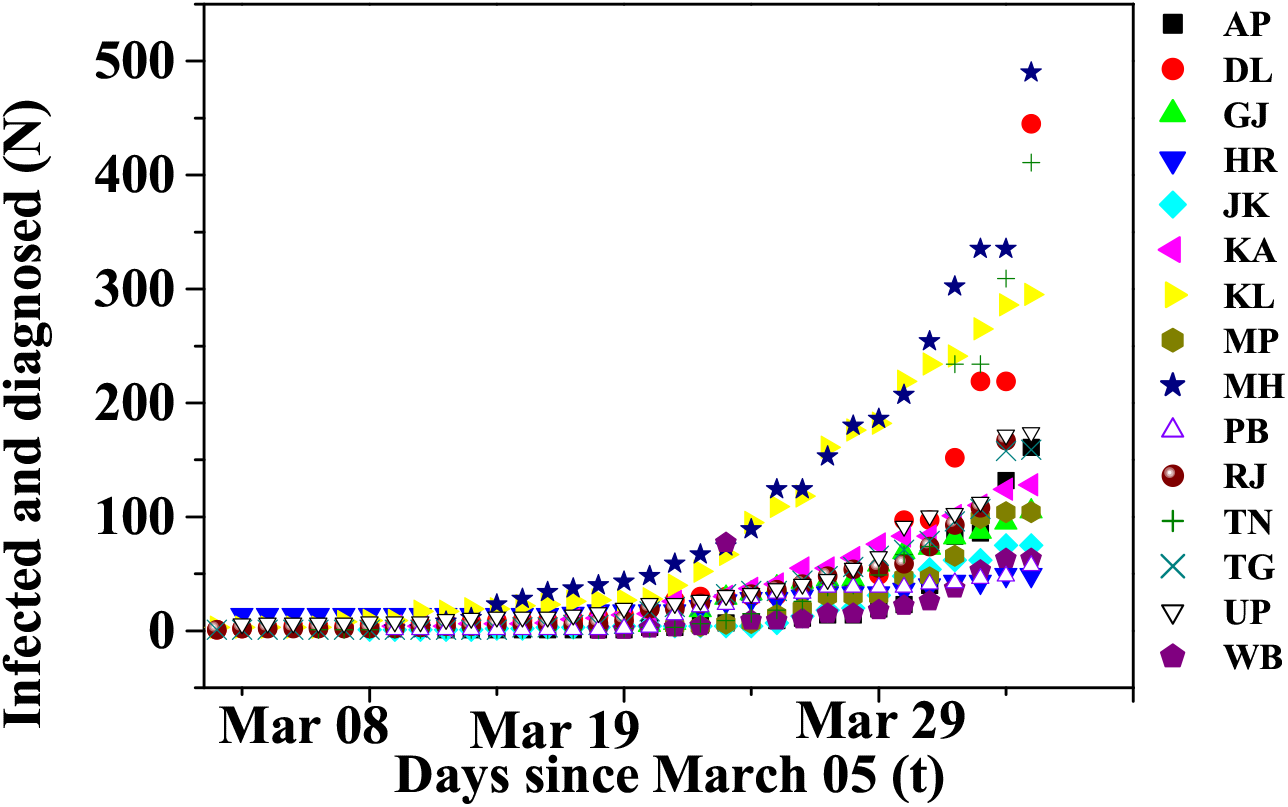
N vs. t plots for 15 states and union territories in India. AP:Andhra Pradesh; DL: Delhi; GJ: Gujarat; HR: Haryana; JK: Jammu Kashmir; KA: Karnataka; KL: Kerala; MP: Madhya Pradesh; MH: Maharashtra; PB: Punjab; RJ: Rajasthan; TN: Tamil Nadu; TG: Telengana; UP: Uttar Pradesh; WB: West Bengal

1. As mentioned earlier, for Italy and China, *N*(*t*) varies exponentially with *t*. To check whether we find similar exponential rise in the number of infected individuals in India, we tried to fit the data points (*N*(*t*)) for each state using an exponential function of *t*. Figure 2 provides a magnified view of the best fitted curves along with the data points for some of the states, using an exponential function, as in Eqn. 1. The full plot for WB is shown in the inset. The increase in *N*(*t*) in neither of the six states, as shown in Figure 2 follows an exponential growth w.r.t. *t* till April 10, 2020. A similar observation is noted and can be made for other states too.
2. For all states there is a hump before a sharp rise in *N*(*t*), which we show in the orange shaded regions in Figure 3, for some of the states. We observe the same feature for other states too. In fact, for some of the states we observe two humps before the sharp rise, as in case of GJ. Interestingly, such a hump prior to a steady and sharp rise can be seen in *N*(*t*) vs. *t* plots for China or Italy. The existence of this hump seems to tally with the well-known model in epidemiology, known as the SIR model [18]. The exact solution of this model [19], [20] is known to show a hump like behaviour when the number of infected individuals is plotted against time. The reason behind why such a hump precedes a sharp rise seems unknown.
3. The data points (*N*(*t*)) for all states beyond the hump can be best fitted by a linear relation, *N*(*t*) = *γ* + *αt*, where *α* is the slope of the plots, determining the characteristics of the rise in the number of infected individuals in a state.

**FIG. 2.**
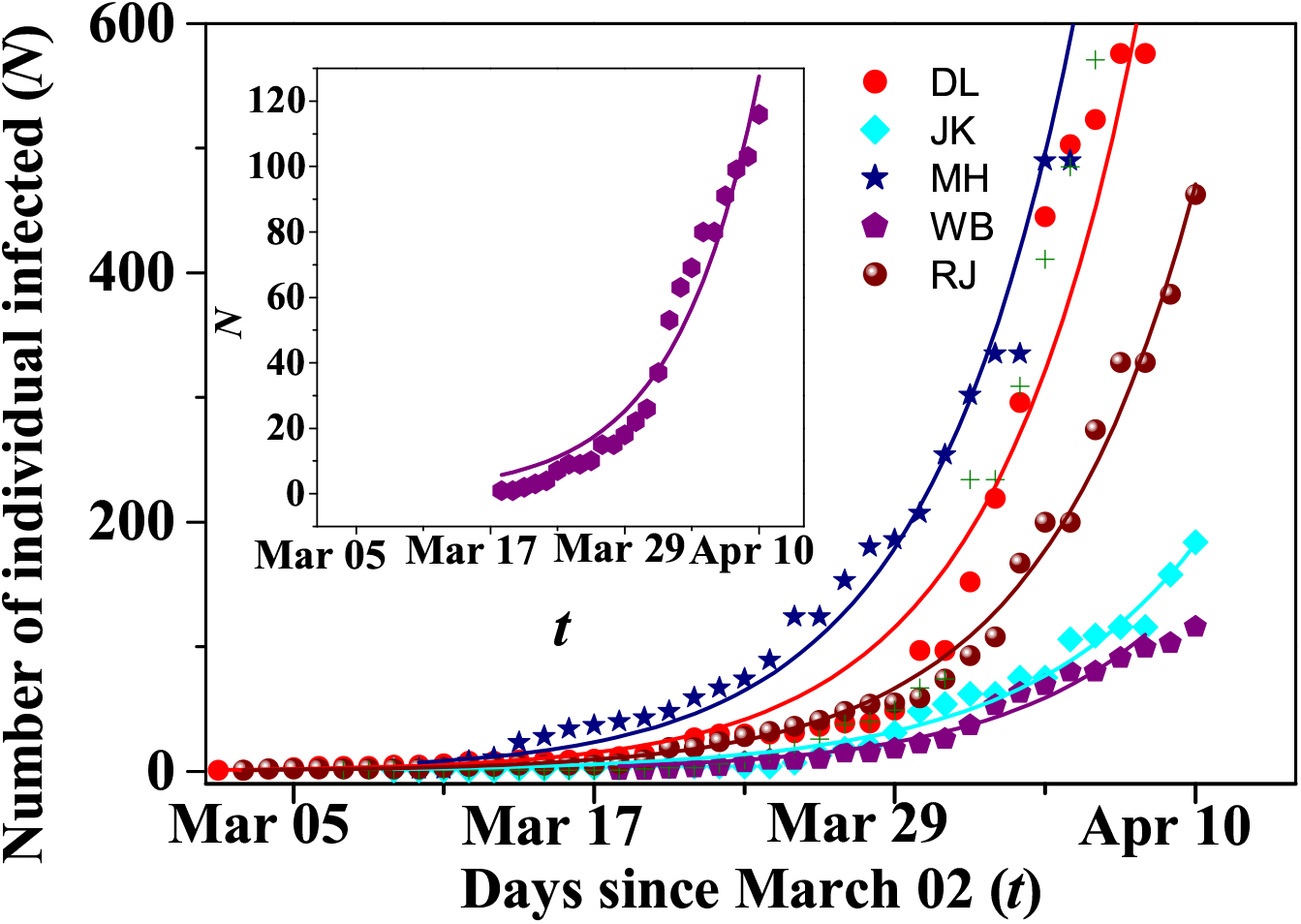
Magnified view of the exponential fit to data points for some states. The full plot for WB is shown in the inset.

**FIG. 3.**
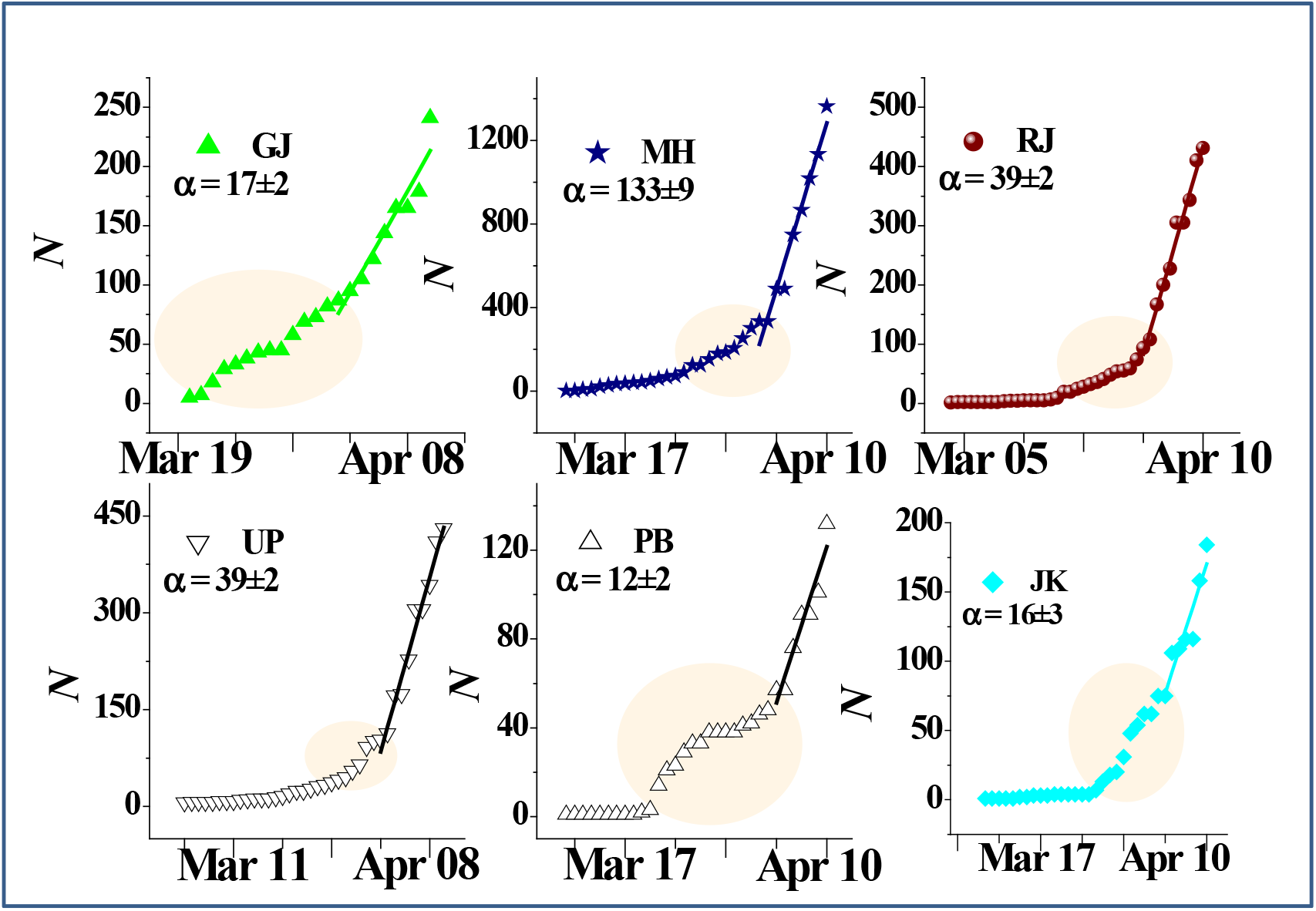
The plot showing the hump and the straight line fit at later dates for some states.

Thus, from the above *N*(*t*) versus *t* plots one can extract the slopes related to the linear growth of the number of infected individuals in each state. We will now work with these slopes and try to correlate them with the temperature, humidity and population density in each state.

As an aside, following a *tanh* model proposed by Zullo [21], we fit *N*(*t*) vs. *t*, for the state KL using the relation *N*(*t*) = *A* + *B* tanh(*βt*) + *δ*. The fitted curve is shown by a red solid line in Figure 4. For most of the other states, it is too early for N(t) to reach the saturation level, and hence it is not meaningful to use this functional form to fit the data for these states. Interestingly, for KL, things do seem to match with this approximate model.

**FIG. 4.**
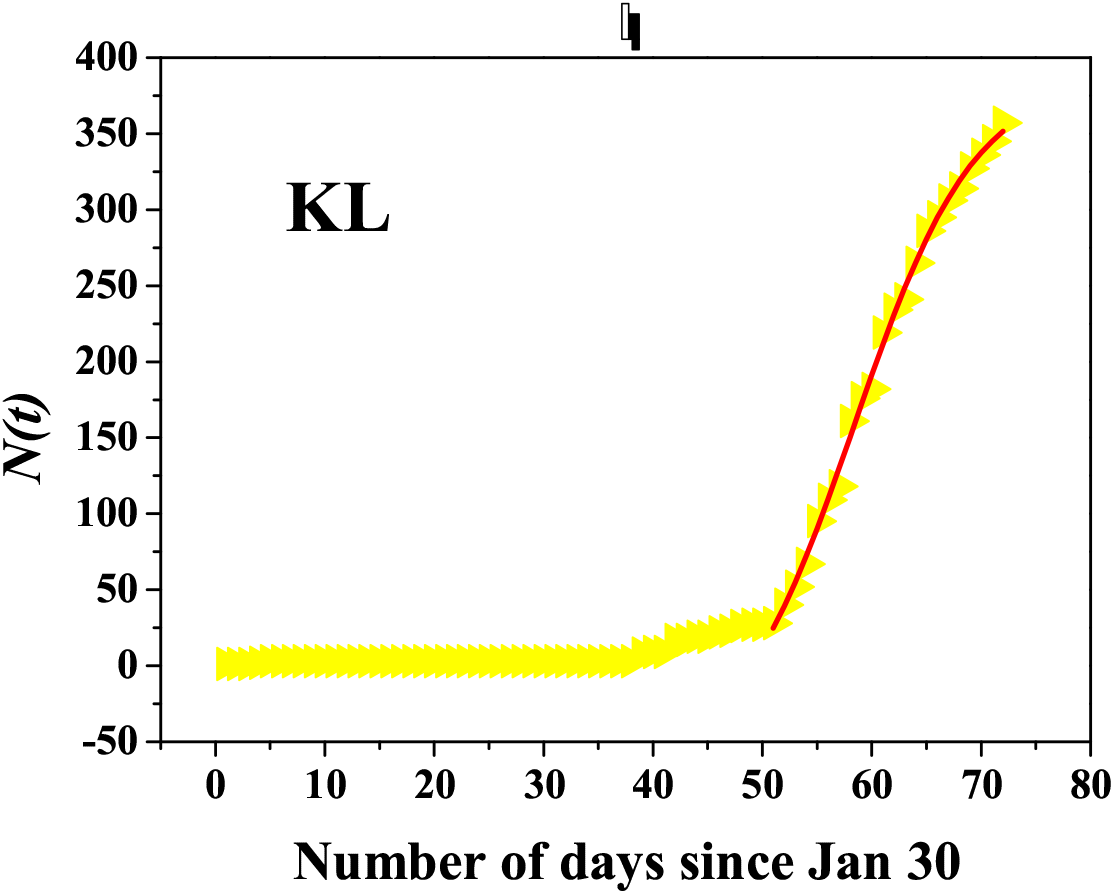
Plot of *N*(*t*) for the entire time frame in Kerala. The red solid line is the best fit to the data points, with the function tanh(*βt*)

## III. VARIATION OF SLOPE WITH AVERAGE MAXIMUM/MINUMUM TEMPERATURES, HUMIDITY AND POPULATION DENSITY

As noted above, for all states, the sharp rise beyond the hump, till April 10, 2020 follows a nearly linear relation with the number of days, and could be fitted fairly well by a straight line, as shown more distinctly, for a few states in Figure 3. Next, we try to find if there is any correlation between the estimated slope and the mean maximum and minimum temperatures (*T*_*max*_, *T*_*min*_) and humidity (*h*) of the states during this growth period. We have estimated the value of *T*_*max*_ and *T*_*min*_ of each state by taking the average of all corresponding recorded temperatures over the days in March-April when the first case was reported and till April 10, 2020. For example, in case of WB, the average of all maximum and minimum temperatures of all days from March 18 to April 10, 2020 are considered to estimate *T*_*max*_ and *T*_*min*_. The data source for the weather parameters is the web portal Accuweather [22] and we note temperatures for the capital of each state (eg. Kolkata for WB). This, we believe, is not a bad approximation, as most of the Covid-19 cases are reported from the capital cities, rather than from remote villages. The variation of the slope with the *T*_*max*_ and *T*_*min*_ and the expected humidity (*h*) for the states over the rise period are shown in Figure 5(a)-(c), respectively. Figure 5(d) plots the variation of the slope with the population density (*ρ*) of the states as per Census 2011 [23]. The data (on population density) is available in the web portal of the Office of the Registrar General and Census Commissioner, India, Ministry of Home Affairs, Government of India.

**FIG. 5.**
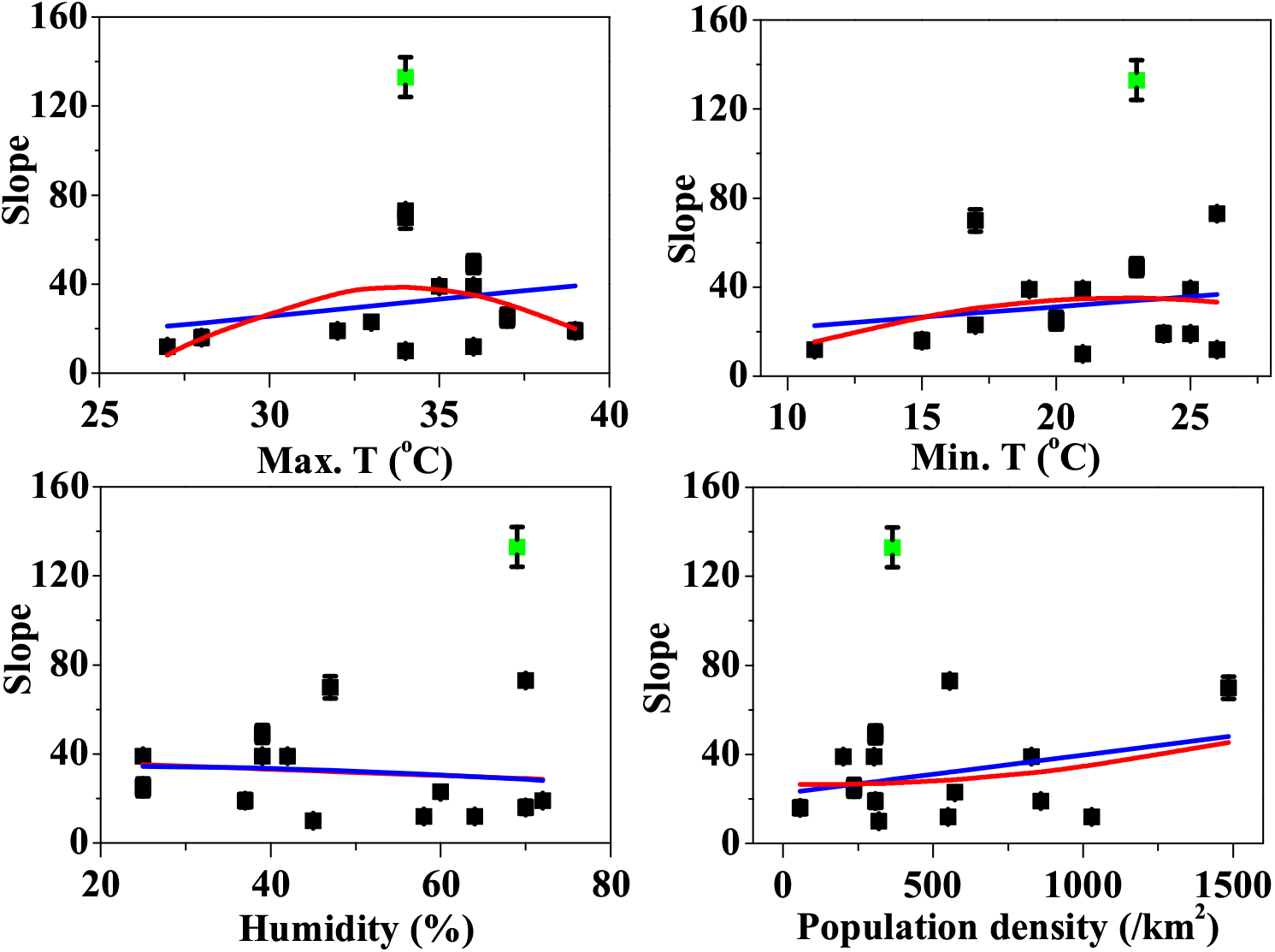
Variation of the slope with (a)maximum temperature, (b) minimum temperature, (c) humidity and (d) population density of the states.

Figure 5 shows that there is no *strong correlation* between the above mentioned parameters and the degree of rise (slope, *α*) of the number of infected cases. Similar observations are made for other countries too, in [1]. Nonetheless, *weak correlation* between the meteorological parameters (*T*_*max*_, *T*_*min*_ and *h*) and the population density (*ρ*) do exist. In view of this, we fitted the data points in each panel in Figure 5 with a linear relation, *α* = *A* + *B* × *x* and with a polynomial relation *α* = *A* + *B* × *x* + *C* × *x*^2^, with *x* as *T*_*max*_, *T*_*min*_, *h* or *ρ*. The best fit curves are shown by blue and red solid lines in Figure 5(a)-(d), respectively. The degree of correlation (*R*^2^), as obtained from the fitting procedures in each panel is shown in Table 1. We find a weak correlation between the value of *α* and the parameters *x*. The value of *R*^2^ is relatively higher if one assumes polynomial relations between the slope (*α*) and *T*_*max*_, *ρ* than if we consider a linear relation. In the fitting procedure, the data point for Maharashtra (shown by green symbol in each plot) is not included, as it lies much away from other data points.

**TABLE 1.** Values of *R*^2^ for the various parameters in the *α*(slope) vs. parameter (*T*_*max*_,*T*_*min*_, *ρ, h*) plots in Figure 5. The value of *R*^2^=1 for perfect correlation.

| Qty. | $R^2$ (polynomial) | $R^2$(linear) | Comment |
| --- | --- | --- | --- |
| $T_{max}$ | 0.22 | 0.06 | Weak correlation (polynomial) |
| $T_{min}$ | 0.06 | 0.04 | No correlation |
| $\rho$ | 0.15 | 0.10 | Very weak correlation |
| $h$ | 0.01 | 0.01 | No correlation |

## IV. RESULTS AND CONCLUSIONS

Our analysis above lead to the following simple conclusions.

- The growth of infected individuals in India, as of now, is nowhere near an exponential rise. The nature is at best a linear growth. This is expected to saturate and at the saturation point the turnaround may be fitted with a *tanh* function (as evident in the plot shown for the state of Kerala). Note that the slopes for each state is different. Is that a hint that climate or other factors may play a role? Without more data any definitive answer is virtually impossible.
- The plot of the slopes w.r.t. the maximum temperature in each state capital during the month of March-April show a weak correlation with *T*_*max*_. The slope seems to follow a polynomial relation with maximum temperature, with an initial rise, a maximum and then a later fall-off. It turns out that the slope is nearly independent of minimum temperature and humidity and the correlation is negligible. We also find a weak linear relation with the population density with a somewhat better correlation as compared to *T*_*min*_ and *h*.
- It may be argued that during the month of March, variation of temperature and humidity across states is not much. However, even within this limited range of values a weak correlation between *α* and *T*_*max*_ could be noted. Whether a weak correlation is just because of less available data or is an actual biological feature of the virus, remains to be seen, experimented upon and understood. At the very least, as apparent from and our very modest and limited analysis and the work by Notari [1], one cannot possibly deny or support a role of climatological factors in the transmission and sustenance of Covid-19.

Keeping in mind that our analysis is based on a large number of assumptions, we would like to end with some comments.

- It is apparent that population density plays a role in the ultimate effects which are expected when a lock-down is imposed in a state. Say, in two states 1 and 2, the number of people in any unit area are *ρ*_1_ and *ρ*_2_, respectively. If *ρ*_1_ > *ρ*_2_, without any restriction on movement, it is probable that more number of people may come near each other, over a period of time, for state 1, than they may for state 2. It appears from Figure 5(d) that the rise in Covid-19 cases is somewhat independent (a very small positive slope, if any and very weak correlation as shown in Table 1) of whether *ρ*_1_ is greater or less than *ρ*_2_. Thus, it is necessary to further investigate whether the ongoing lock-down (which is within the period over which we have analysed the data here) is the cause behind this feature of the slope versus population density plot in Figure 5 (d)?
- It is highly non-trivial to propose a universal model to describe a pandemic. The habits and living styles of the vast majority of a community surely play a big role in spreading the disease. It becomes quite subjective and artificial if we try to quantify such behavorial features. For example, the data point for MH, which includes Mumbai, a cosmopolitan business city in India, is an outlier in the plots, as shown in Figure 5. One is not sure why–is it because of more testing in Mumbai, non-implementation of rigorous lock-down, unknown environmental factors – one may never know the answers to such questions, in terms of numbers.
- In the next few days, if the growth of *N*(*t*) shows trends towards an exponential rise one could still believe in the *α* found here. *α* would then become the exponent in the exponential, but we all know that for small *t* the exponential behaves linearly. In the event of an exponential rise, the data used here would be for the small-*t* time-frame with *α* as the slope in this initial phase of a linear rise in *N*(*t*).

Finally, it will be the precise and robust experimental results from biological/medical research which can eventually shed more light on how to control such a pandemic. Epidemiological models, which certainly rely on such precise biological/medical inputs can only give good qualitative hints, but may not necessarily be the best way of predicting the future.

## Data Availability

Data used are freely available in the internet. References have been provided in the manuscript.

## Endnote

**As and when newer data becomes available, we hope to modify the results quoted in this work here**.

## References

[1] A. Notari, Temperature dependence of COVID-19 transmission, 2003:12417.

[2] K. H. Chan, J. S. Malik Peires et al. The effects of temperature and relative humidity on the viability of the SARS Coronavirus, Advances in virology, Vol. 2011 (Article ID 734690), 2011.

[3] M. Wang, A. Jiang et al. Temperature significant change COVID-19 transmission in 429 cities, www.medrxiv.org 20025791 (2020).

[4] M. Araujo and B. Naimi, Spread of SARS-Cov-2 Coronavirus likely to be constrained by climate, www.medrxiv.org 20034728 (2020).

[5] Q. Bukhari and Y. Jameel, Will coronavirus pandemic diminish by summer?, available at ssrn.com/abstract=3556998 (2020).

[6] K. F. Weifeng, Lv Jingyuan Wang, Ke Tang, High Temperature and High Humidity Reduce the Transmission of COVID-19, arxiv:2003.05003 [q-bio.pe]. 2020.

[7] M. Sajadi, P. Habibzadeh, A. Vintzileos, S. Shokouhi, F. Miralles-Wilhelm and Anthony Amoroso, Temperature, Humidity and Latitude Analysis to Predict Potential Spread and Seasonality for COVID-19, Available at ssrn.com/abstract=3550308 or dx.doi.org/10.2139/ssrn.3550308.

[8] W. Luo, M. S Majumder, D. Liu, C. Poirier, K. D Mandl, M. Lipsitch, and M. Santillana, The role of absolute humidity on transmission rates of the COVID-19 outbreak, www.medrxiv.org, 20022467 (2020)

[9] A. Brandenburg, Quadratic growth during the 2019 novel coronavirus epidemic, arxiv:2002.03638 (2020).

[10] R. Ranjan, Prediction for Covid-19 outbreak in India using epidemiological models. arxiv: www.medrxiv.org 200451466 (2020)

[11] A. Senapati, S. Rana, T. Das, J. Chattopadhyay, Impact of intervention on the spread of Covid19 in India: a model based study, 2004.04950 (2020).

[12] T. Sardar, S. Nadim and J. Chattopadhyay, Assessment of 21 days lockdown effect in some states and overall India: a predictive mathematical study on COVID-19 outbreak, 2004.03487 (2020).

[13] S. Das, P. Ghosh, B. Sen and I. Mukhopadhyay, Critical community size for COVID-19 - a model based approach to provide a rationale behind the lockdown, 2004.03126 (2020).

[14] R. Singh and R. Adhikari, Age-structured impact of social distancing on the COVID-19 epidemic in India, 2003:12055 (2020).

[15] S. Gupta and R. Shankar, Estimating the number of COVID-19 infections in Indian hot-spots using fatality data, 2004.04025 (2020).

[16] K. Biswas and P. Sen, Space-time dependence of corona virus (COVID-19) outbreak, 2003.03149 (2020).

[17] Data taken from : www.mohfw.gov.in/. Also available at en.wikipedia.org/wiki/2020 coronavirus pandemic in India.

[18] The SIR model and its variants like the currently used SEIR model with and without social distancing factors has been around since 1932-33 since the seminal work of W. O. Kermack and A. G. McKendrick, Contributions to the mathematical theory of epidemics, Proc. R. Soc. Lond. A 138, 55(1932), ibid bf A 141, 94 (1933). See also the book by D. J. Daley and J. Gani, Epidemic Modeling: an introduction, Cambridge University Press, Cambridge (2005).

[19] T. Harko, F. S. N. Lobo and M. Mak, Exact analytical solutions of the Susceptiple-Infected-Recovered (SIR) epidemic model and the SIR model with equal death and birth rates, Applied Mathematics and Computation 236, 184 (2014).

[20] M. Cadoni, How to reduce epidemic peaks keeping under control the time-span of the epidemic, 2004.02189.

[21] F. Zullo, Some numerical observations about the COVID19 epidemic in Italy 2003.11363.

[22] Data from the website www.accuweather.com

[23] Census 2011 details can be found in the site: en.wikipedia.org/wiki/List_of_states_and_union_territories_of_India_by_population

